# Long-term performance of intracortical microelectrode arrays in 14 BrainGate clinical trial participants

**DOI:** 10.1101/2025.07.02.25330310

**Authors:** Nick V. Hahn, Elias Stein, BrainGate Consortium, John P. Donoghue, John D. Simeral, Leigh R. Hochberg, Francis R. Willett

## Abstract

Brain–computer interfaces have enabled people with paralysis to control computer cursors, operate prosthetic limbs, and communicate through handwriting, speech, and typing. Most high-performance demonstrations have used silicon microelectrode “Utah” arrays to record brain activity at single neuron resolution. However, reports so far have typically been limited to one or two individuals, with no systematic assessment of the longevity, decoding accuracy, and day-to-day stability properties of chronically implanted Utah arrays. Here, we present a comprehensive evaluation of 20 years of neural data from the BrainGate and BrainGate2 pilot clinical trials. This dataset spans 2,319 recording sessions and 20 arrays from the first 14 participants in these trials. On average, arrays successfully recorded neural spiking waveforms on 35.6% of electrodes, with only a 7% decline over the study enrollment period (up to 7.6 years, with a mean of 2.8 years). We assessed movement intention decoding performance using a “decoding signal-to-noise ratio” (dSNR) metric, and found that 11 of 14 arrays provided meaningful movement decoding throughout study enrollment (dSNR > 1). Three arrays reached a peak dSNR greater than 4.5, approaching that achieved during able-bodied computer mouse control (6.29). We also found that dSNR increases logarithmically with the number of electrodes, providing a pathway for scaling performance. Longevity and reliability of Utah array recordings in this study were better than in prior nonhuman primate studies. However, achieving peak performance consistently will require addressing unknown sources of variability.

## Introduction

Brain-computer interfaces have emerged as a promising approach for restoring movement and communication to people with paralysis. Successful demonstrations have included point-and-click cursor control (Jarosiewicz et al. 2015; Pandarinath et al. 2017), robot control (Hochberg et al. 2012; Collinger et al. 2013; Benabid et al. 2019), functional electrical stimulation (Ajiboye et al. 2017), typing (Willett et al. 2021; Vansteensel et al. 2016, 2024; Shah et al. 2024; Jude et al. 2025), and speech (Moses et al. 2021; Willett et al. 2023; Metzger et al. 2023; Card et al. 2024; Wairagkar et al. 2025). Many of the highest performing demonstrations to date have used silicon microelectrode arrays (the “Utah” array) to record brain activity at single neuron resolution (Collinger et al. 2013; Pandarinath et al. 2017; Willett et al. 2021; Card et al. 2024; Wairagkar et al. 2025), providing rich information about the user’s motor intent.

However, nearly all reports in humans have been limited to case studies of one or two participants, including those that have focused on summarizing array recording performance over time (Simeral et al. 2011; Downey et al. 2018; Sponheim et al. 2021; Colachis et al. 2021; Hughes et al. 2021). Key questions remain about how decoding accuracy varies across individuals, how long neural recordings last over years, and how stable the recordings are across days. Answers to these questions are needed to place isolated demonstrations into context and understand whether they represent typical signal quality. A more comprehensive evaluation of the Utah array can also set a benchmark against which improvements can be measured, informing the design of future recording devices.

Here, we present a post-hoc analysis of neural recording and motor decoding performance of the Utah array in the first 14 participants enrolled in the BrainGate and BrainGate2 pilot clinical trials between 2004 and 2019 (constituting all available participants at the time when this analysis was started, but not including more recent participants enrolled between 2021 and 2025). Encouraging safety results from these same 14 participants were recently reported in an interim study (Rubin et al. 2023). Like prior assessments in nonhuman primates (Suner et al. 2005; Chestek et al. 2011; Barrese et al. 2013; Sponheim et al. 2021), here we evaluated the percentage of electrodes which successfully recorded neural spiking waveforms over time (array “yield”). Additionally, since most arrays were placed in the same brain region (hand knob area of motor cortex) and all participants regularly engaged in BCI-enabled cursor control tasks, we were able to compare neural decoding quality across the 9 participants for which high-bandwidth (30 kHz) neural data was available. We assessed the stability of neural tuning over time, how cursor decoding accuracy compares to mouse movement accuracy in able-bodied volunteers, and how decoding accuracy scales with the number of electrodes (which is important for understanding how higher channel count devices might improve decoding). These functional measurements are more directly relevant for BCI applications than array yield, particularly because low-amplitude background activity can be highly useful for neural decoding even if it fails to generate clear spiking waveforms (Nason et al. 2020).

## Results

Between 2004 and 2019, 14 participants were enrolled in the BrainGate and BrainGate2 pilot clinical trials; each participant had one or two 96-channel platinum-metalized Utah microelectrode arrays placed in cortex. Nineteen of the total twenty arrays were placed in the hand area of motor cortex (Figure 1A, B), with one array placed in the middle frontal gyrus. The BrainGate and BrainGate2 clinical trials were designed to evaluate the safety of implanted microelectrode arrays and to discover fundamental insights into the effective design of intracortical BCIs. Testing new neural decoding architectures and task designs was common throughout the trials, and as such it is not feasible to summarize across all types of data collected.

**Fig. 1.**
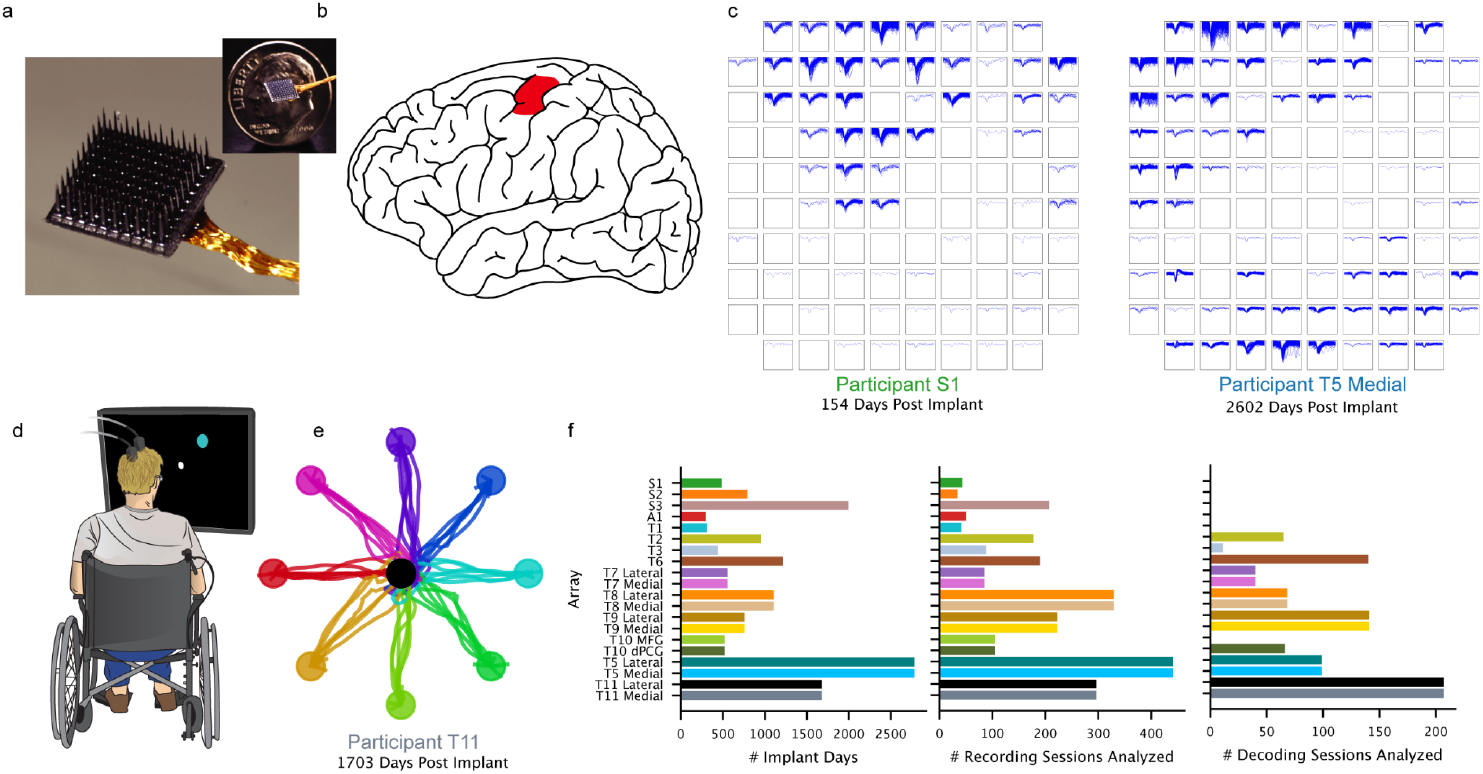
Datasets curated for assessing recording and decoding performance. **(A)** One or two 96 channel intracortical microelectrode arrays were placed in the cortex of 14 participants as part of the BrainGate and BrainGate2 clinical trials. 19 of 20 arrays were placed in the hand knob area of the dominant precentral gyrus **(B)**, and one was placed in the middle frontal gyrus of Participant T10. **(C)** Example spike waveforms. For this study, 30 Khz recordings were decimated to 15 Khz, band pass filtered with a pass band of 250-5000 Hz, re-referenced with linear regression referencing (LRR) and thresholded at −4.5 times the robust standard deviation of the voltage signal for each channel. **(D)** To assess decoding performance, we retrospectively analyzed sessions in which participants were asked to attempt to move a computer cursor towards a defined target. Imagery varied between sessions and participants. Tasks included Fitts, Radial-8, grid tasks and their variations. To assess array recording yields, data from all types of sessions were used (including sessions where no cursor control occurred). **(E)** Example cursor trajectories for a Radial-8 task. **(F)** Overview of recording duration and sessions analyzed. Total array implant days ranged from 296 to 2780 with a mean of 1063. After excluding sessions with excessive electrical noise, 2,319 recording sessions and 634 decoding sessions were analyzed. T10’s middle frontal gyrus array was only used for recording evaluation, as it was in a different brain area with neural tuning properties not comparable to the hand knob area. High bandwidth neural data (30 kilo-samples per second) required for retrospectively evaluating decoding performance in a uniform manner were not consistently available for participants S1, S2, S3, A1, and T1, and so these participants were not included in decoding analyses.

To summarize findings spanning 14 participants over 15 years, here we curated two types of datasets: (1) a “yield dataset” containing a single neural recording of at least 5 minutes duration from every research session, which was used to estimate array yield (Figure 1D), and (2) a “decoding dataset” consisting of all neural recordings during which a BCI was used to move a computer cursor towards well-defined targets (Figure 1E), which was used to estimate neural decoding accuracy, neural tuning stability and electrode count scaling properties. This dataset consists of a more limited number of days and participants, because not every experimental session included BCI cursor control, and because for the first 5 participants high bandwidth neural data (30 kHz) was either not recorded during cursor control (S1, S2, A1), not recorded for the first year post-implant (S3), or could not be consistently aligned to cursor task data (T1).

Note that prior publications have shown successful demonstrations of neural cursor control in S1 and S2 (Hochberg et al. 2006), S3 (Kim et al. 2008; Simeral et al. 2011; Jarosiewicz et al. 2013; Bacher et al. 2015; Jarosiewicz et al. 2015), A1 (Kim et al. 2008), and T1 (Jarosiewicz et al. 2013).

### Neural spiking activity persists for multiple years with little average decline

We first investigated how the number of electrodes able to detect spiking activity (“yield”) changed over time, looking across a total of 2,319 recording sessions. Spiking activity was defined as the detection of at least 2 spiking events per second when using a threshold of −4.5 times the robust standard deviation of the voltage signal (see Methods). We observed substantial variability across 20 arrays in 14 different participants (Figure 2A), with some arrays sharply declining in yield within the first year (S2, S3 and T2) or having consistently low yield (T3), some arrays increasing over time (T7, T8), and many arrays staying relatively constant or gradually declining over time (e.g., T5, T6, T10, T11). Yield trajectories in the first year were not always predictive of yields over longer periods (e.g., T7’s medial array increased in yield after the first year). Yield curves were estimated using locally weighted scatterplot smoothing (LOWESS)(Cleveland 1979). Although the cause of S2 and T2’s sudden declines in yield are not known, S3’s decline coincided with and may have been triggered by mechanical trauma to the pedestal that occurred outside of research study sessions. T3’s array, which had a consistently very low yield, was believed to have had its wire bundle unintentionally placed under tension during surgery, which could have resulted in the recording tips being largely epicortical rather than intracortical.

**Fig. 2.**
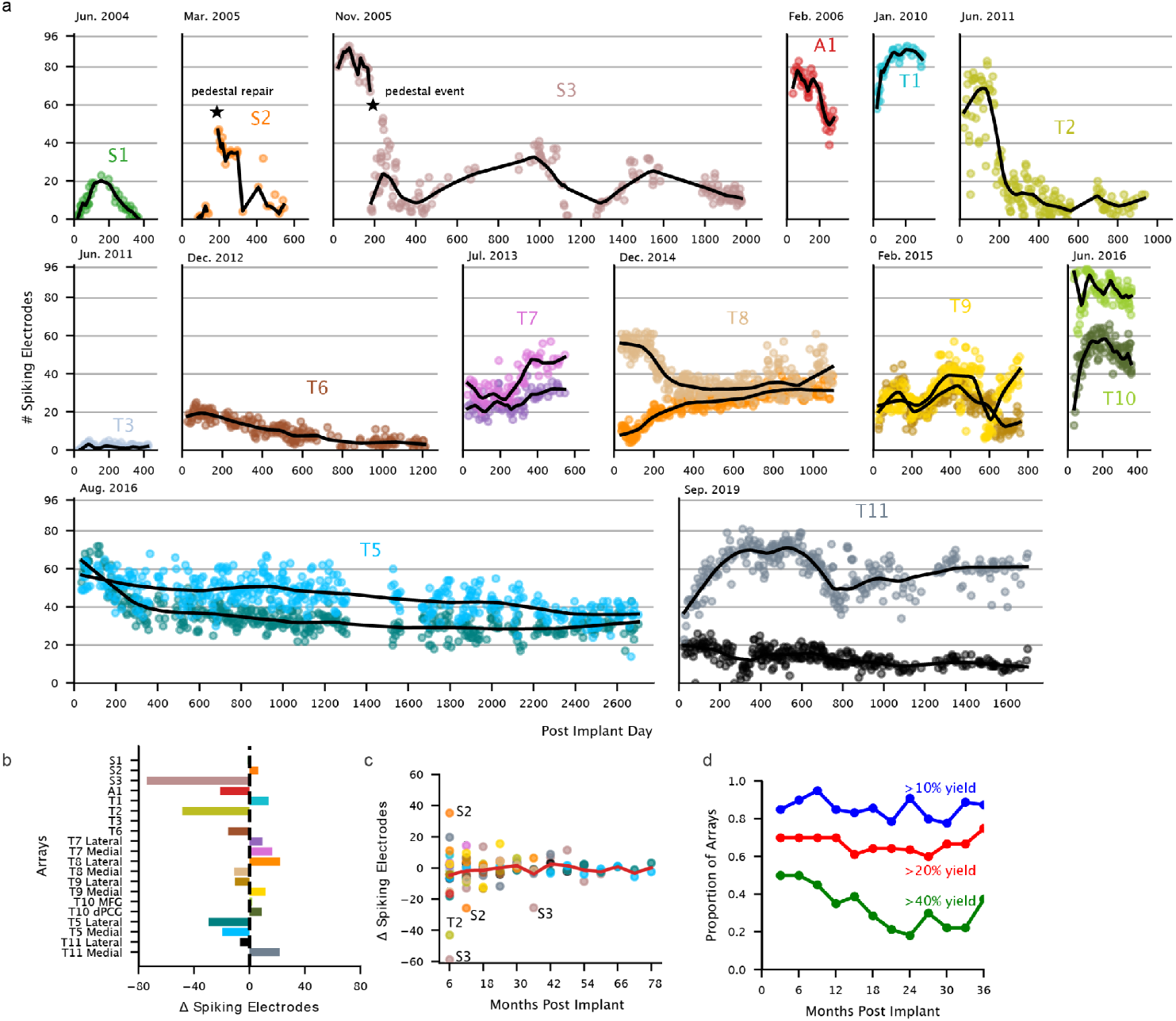
Long-term trends in neural spiking activity recording performance. **(A)** Array yield over the recording duration of twenty 96-channel microelectrode arrays. Each point represents the number of electrodes detecting spiking activity in a single neural recording of 5 minutes duration or longer (one recording per session). Spiking electrodes are defined as electrodes detecting a threshold crossing rate greater than 2 Hz when using a −4.5 x robust standard deviation threshold (see Methods). LOWESS curves are shown in black. For participants with two arrays, data points in the lighter shade represent the more medial array. Implant date is indicated above each participant panel. Participant S2’s array experienced a steep increase in yield after repair of the percutaneous pedestal (day 190) which placed conductive epoxy on each bond site to restore connections. Months later, array yield declined sharply again (day 323), and it is unknown whether this was due to failure of the previous repair or an unrelated event. Participant S3 experienced an accidental forceful contact of the pedestal against her headboard on the evening of day 181, with a decrease in yield that slowly, partially recovered. **(B)** Overall deltas in yield. Deltas were calculated by taking the mean number of spiking electrodes in sessions recorded during the last 3 months post implant and subtracting the mean number of spiking electrodes recorded during the first 3 months of recording. **(C)** 6 month deltas in yield, up to six and a half years post-implant. Each point represents the mean spiking electrode count over a 6 month period minus the mean spiking electrode count over the previous six month period. The red line plots the mean of deltas. Outliers are labeled, with participant T2, S2, and S3 all experiencing a steep decline. **(D)** Array proportions exceeding a certain yield over the first 3 years of recording. If data was not recorded from an array during a certain time period, that array was not included in the calculation for that time period.

Averaged over all days, participants, and arrays, mean array yield (percentage of electrodes recording spiking activity) was 35.6%. Arrays experienced only a modest 7% average decline in yield between the first 3 months of recording and the last three months of recording (Figure 2B; 41% yield in the first three months to 34% yield in the last three months). However, there was large variability in yield across arrays (standard deviation: 20.5%). A minority of arrays in a subset of participants contributed substantially to this variability, including the three arrays that showed a sharp decrease in yield over the first year (S2, S3, and T2; Figure 2C). Overall, results were favorable compared to prior work in nonhuman primates, with over 50% of arrays maintaining at least 20% yield over the first three years (Figure 2D; compare to Sponheim et al. 2021, where less than 15% of arrays maintained a 20% yield). This demonstrates the potential of long-term, reliable detection of neural spike waveforms with planar microelectrode arrays in humans.

We also examined electrode impedance and found that it increased sharply post-implant, followed by a consistent and gradual decline (Supplementary Figure 1) consistent with prior reports (Barrese et al. 2013; Barrese, Aceros, and Donoghue 2016). Spike waveform amplitude and background noise amplitude also declined over time (Supplementary Figure 2A-B), and smaller waveform amplitudes were associated with lower impedance values (Supplementary Figure 2C). Since both the spike waveform and background noise amplitudes declined at similar rates, the waveform signal-to-noise ratio remained relatively constant over time (Supplementary Figure 2D). This overall decline in signal amplitude and impedance may be due to insulation degradation (Barrese et al. 2013; Barrese, Aceros, and Donoghue 2016), particularly around the edges of the planar array where low impedances were sometimes observed progressively over time (e.g., T2, T3, T5, T6; Supplementary Figure 3). Nevertheless, these changes did not appear to affect detection of spiking signals or decoding performance (see next section).

### Movement intention can be decoded reliably for years in most arrays

Next, we examined the extent to which cursor movement intention could reliably be decoded from intracortical recordings, how decoding varied across arrays, and how it compared to the accuracy of able-bodied cursor control using a computer mouse. A total of 634 sessions across 14 arrays and 9 participants were analyzed to evaluate movement intention decoding during closed loop cursor control tasks (Figure 1F).

To evaluate the amount of intended movement information in the neural signals recorded by an array, we quantified how well an offline linear decoder could transform neural activity into a “movement intention” unit vector pointing from the cursor to the target, using neural features from a short time window at the beginning of each movement. The decoder output from each trial can be visualized as a dot in 2D space and colored according to the ground truth direction; the more accurate the decoder is, the more the dots should form a tight ring whose colors form an orderly gradient (Figure 3A). Accurate decoder output represents movement intention vectors that point more accurately at the target with a consistent magnitude, and would thus enable higher quality cursor control, given that cursor control algorithms often use linear mappings between neural features and cursor velocity (Kim et al. 2008; Gilja et al. 2012; Willett et al. 2017, Pandarinath, et al. 2017).

**Fig. 3.**
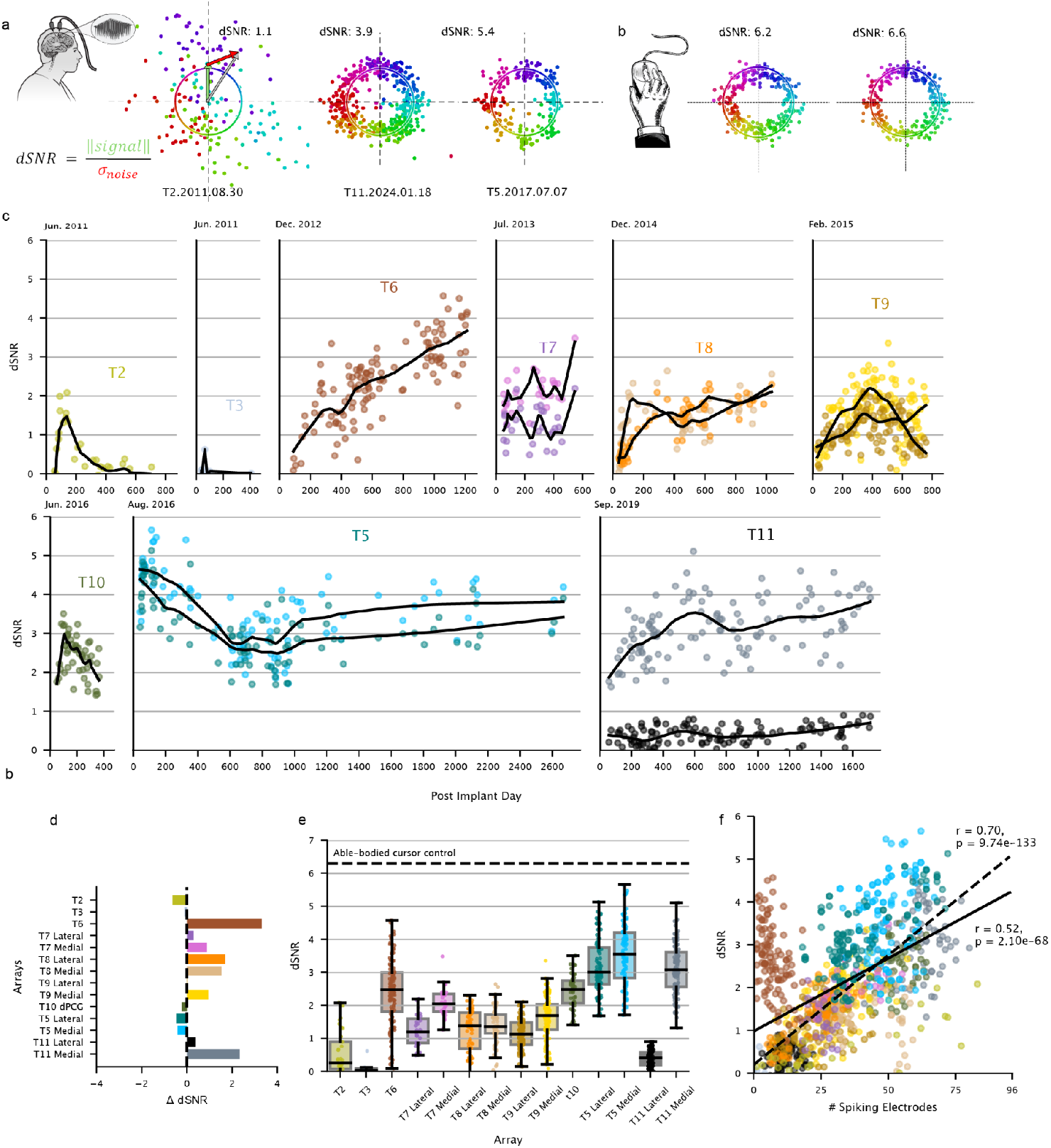
Retrospective assessment of movement decoding signal-to-noise ratio across time. **(A)** Decoding signal-to-noise ratio (dSNR) and example dSNR plots. dSNR is a vector-based, movement intention decoding metric that quantifies how well movement intention can be decoded from all electrodes of a given array during a target-directed cursor control task. To estimate dSNR, retrospectively computed linear decoder output within a 300 ms window is decomposed into a signal component (pointing toward the target) and a noise component to calculate a decoding SNR. Each point in the dSNR plots denotes a trial, color coded by target location. Spike band power neural features were retrospectively computed in a uniform manner for all participants from whom high bandwidth neural recordings were available for analysis. A linear decoder was then trained in a uniform manner to decode these neural features, ensuring that results are independent of whatever algorithms were used at the time of data collection. **(B)** Example SNR plots for able-bodied cursor control using a computer mouse. Cursor trajectories from an initial push towards the target were used to calculate dSNR **(C)** Decoding SNR over time. Each point represents a dSNR value for one session. LOWESS curves are shown in black for each array. **(D)** Overall deltas in dSNR. Deltas were calculated by taking the mean dSNR in sessions recorded during the last 3 months post implant and subtracting the mean of dSNR recorded during the first 3 months. **(E)** Decoding SNR distributions. Arrays are displayed in implant order. The dotted line indicates mean decoding SNR across 9 able-bodied participants using a computer mouse. **(F)** Decoding SNR and array yield are correlated (r = 0.52, *p* = 2e-68). When excluding the T6 array, which had low array yield but high information content in spike band power features, this relationship is more pronounced (r = 0.70, *p* = 9e-133)

To quantify the quality of the offline decoder output, the *decoding signal-to-noise ratio (dSNR)* metric (Willett, Murphy, et al. 2017) decomposes each decoder output vector into a signal and a noise vector (Figure 3A) and takes the ratio of their magnitudes (see Methods). dSNR is independent of the particular task parameters, decoding architecture, and signal processing used in real-time during data collection, and thus enables meaningful post-hoc comparisons across a diversity of datasets that would otherwise not be possible via task performance metrics alone. It can also be used to quantify target-directed able-bodied mouse movements (Figure 3B), contextualizing the neural dSNR values. To compute dSNR for able-bodied mouse movements, we used the endpoint of the computer cursor after an initial “ballistic” center-out cursor movement to define the “decoder output” vector.

There was wide variability in dSNR across participants and arrays (Figure 3C), although on average dSNR increased by 0.34 across the duration of recording (ranging from 1.2 to 7.6 years, Figure 3D). Most arrays maintained dSNR values greater than 1, indicating a persistent ability to decode meaningful movement intention signals. A minority of arrays had low dSNR at the end of first year post-implant and did not recover dSNR over the course of study participation (T2, T3, T11 Lateral). High-performing arrays approached able-bodied dSNR values (Figure 3E; T5 Lateral, T5 Medial, T11 Medial), but typical arrays yielded about 1/3 the dSNR of able-bodied computer mouse movement (average computer mouse dSNR=6.29).

dSNR was correlated with array yield (pearson r: 0.52, *p*: 2.10e-68; Figure 3F), but array yield did not fully account for the large variability of decoding performance, in part due to information contained in “spike band power” features (e.g. participant T6). The results presented above used spike band power features to decode movement intention, since spike band power (power contained in the 250-5000 Hz band) can capture low-amplitude spiking signals that might otherwise be missed by threshold crossing features. We evaluated how dSNR varied as a function of neural features used for decoding, and found that spike band power led to higher performance on average as compared to threshold crossing rates, even at lower thresholds (Supplementary Figure 5). Overall, these outcomes show that long-term persistence of meaningful decoding performance is typical and can occur even at low array yields, although absolute dSNR levels can vary widely across participants and arrays.

### Neural representations of movement intention remain stable on short time scales

Next, we investigated the day-to-day changes in how movement intention is represented in the recorded neural activity. Neural tuning stability was analyzed by using a linear encoding model to describe how spike band power features were tuned to a movement intention vector that points from the cursor to the target. After fitting, model coefficient similarity was assessed by correlating the coefficients of models fit on different days (see Methods). Only arrays from the decoding dataset where movement intention could be decoded reliably (dSNR >1) were analyzed (3 arrays excluded), yielding a total of 593 recording sessions and 11 arrays from 7 participants.

Day-to-day tuning correlations exhibited a variety of patterns (Figure 4A), due in part to potential changes in motor imagery across days and/or tasks, varying task instructions, and variable intervals between consecutive cursor control sessions. Nevertheless, many arrays showed clear regimes of long-lasting tuning stability. Average stability as a function of time elapsed (Figure 4B) was generally high within the time span of 1 month (8 of 11 arrays, r>0.6). This is encouraging, as short-range stability enables decoders calibrated on prior days to be used on future days without requiring user interruption for re-calibration (Card et al. 2024; Wilson et al. 2023; Fan et al. 2023; Hosman et al. 2023).

**Fig. 4.**
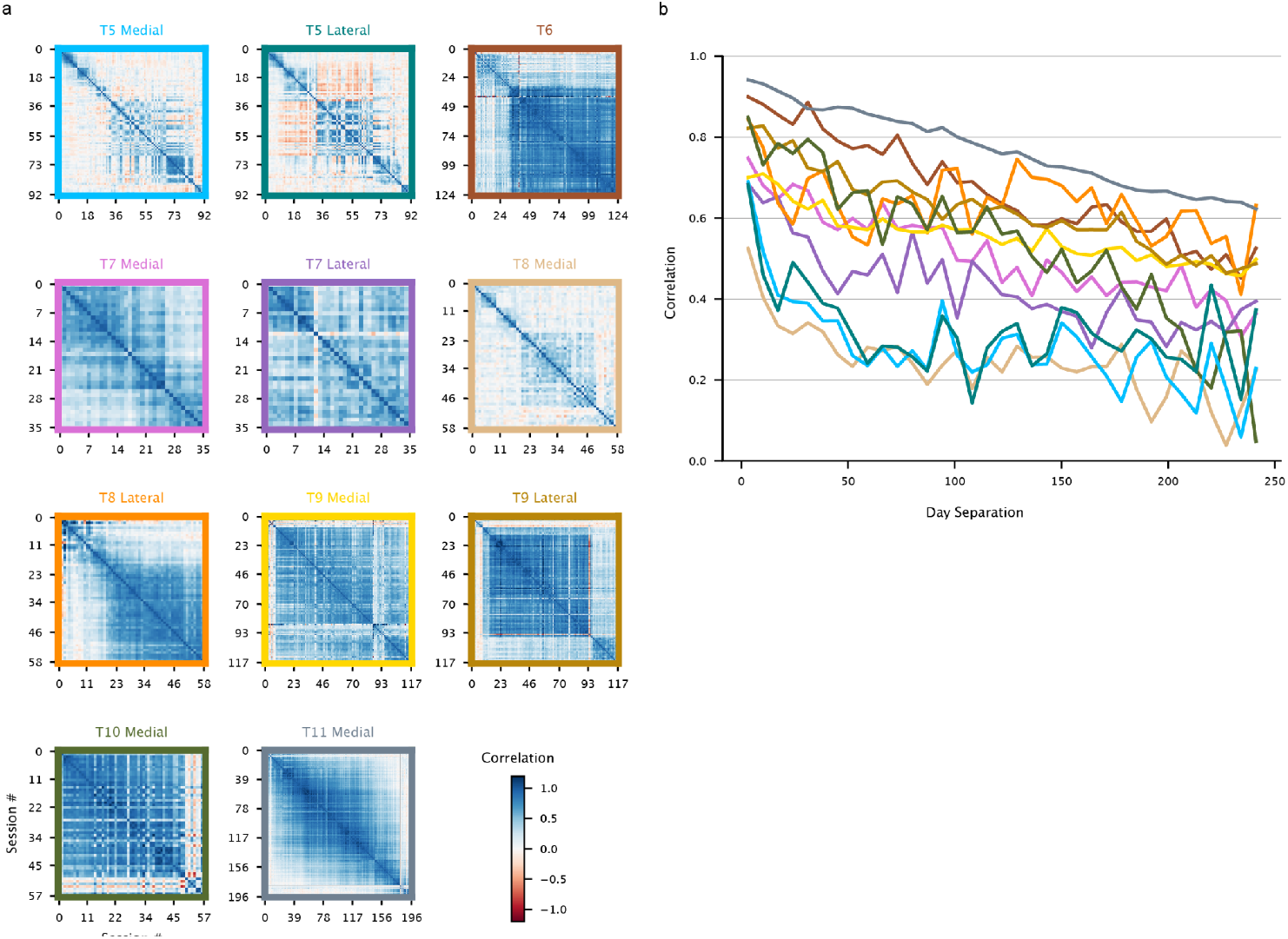
Stability of neural tuning to movement intention across days. **(A)** Tuning stability heatmaps. A linear encoding model was used to describe neural activity as a function of a point-at-target vector that describes the user’s movement intention during a cursor control task. The Pearson correlation between model coefficients for each session pair is shown in each (i,j) entry. Blocks of blue color indicate time periods of high stability. **(B)** Tuning stability as a function of day separation. Tuning remains stable on the time scale of a month (8 of 11 arrays, r>0.6), which could enable decoders calibrated on prior days to be used on future days without requiring re-calibration. Line colors correspond to the heatmap outlines in (a).

### Electrode count scaling provides a pathway to higher performance

Finally, we examined how decoding performance scales with the number of electrodes. We computed average dSNR across all cursor control sessions, for each array and participant, as a function of electrode count by randomly subsampling from all 96 electrodes (n=500 resamplings per electrode count). We found that dSNR scaled as an approximately logarithmic function of the number of electrodes, up to 192 electrodes for participants with dual arrays and 96 electrodes for participants with a single array (Figure 5). Only arrays providing meaningful movement intention decoding (dSNR >1) were evaluated. By linearly extrapolating to 1024 electrodes, a subset of arrays (T5 Lateral, T5 Medial, T6, T11 Medial) demonstrated a projected dSNR that was 70% or greater than the average computer mouse dSNR observed across 9 able-bodied volunteers. These results are supportive of the many ongoing efforts towards higher channel count devices (Musk and Neuralink 2019; Sahasrabuddhe et al. 2021; Hettick et al. 2022), although it appears that for arrays with lower dSNR values (e.g., T8), many orders of magnitude more electrodes would be required to reach able-bodied computer mouse performance.

**Fig. 5.**
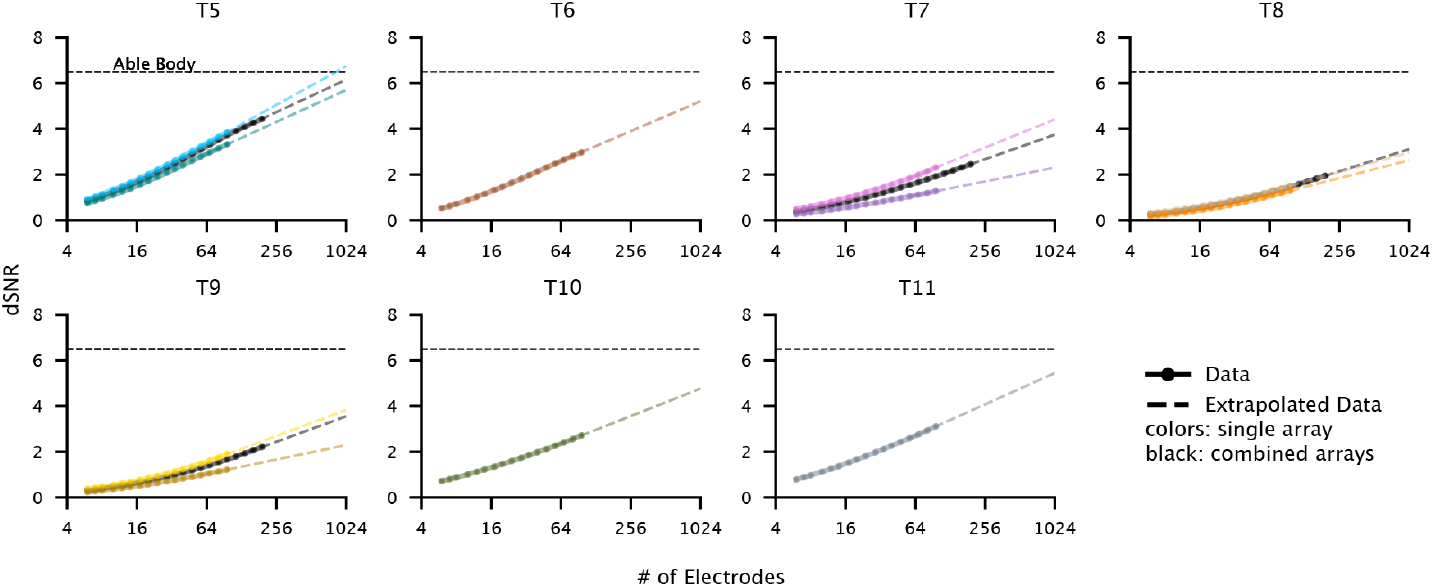
Decoding SNR scales logarithmically with electrode count. For each array, decoding SNR (dSNR) was assessed using a random subset of electrodes to investigate how dSNR varies as a function of electrode count (for each electrode count, dSNR was averaged over 500 random subsets). For each single array, electrode counts ranged from 6 to 96. For participants with dual arrays, combined electrode counts ranged from 6 to 192. Extrapolated data is represented with a dotted line and combined arrays are represented in black. dSNR scales logarithmically with the number of electrodes. Here, for arrays exhibiting higher performance per electrode (e.g., t5, t11), performance approaches able-bodied levels as electrode count is extrapolated up toward 1024 electrodes.

## Discussion

We analyzed the performance of Utah microelectrode arrays placed in the first fourteen participants of the longest running clinical trial for implanted brain-computer interfaces. Overall, our findings show that Utah arrays typically continue to record neural spiking waveforms for years with little average decline, contrary to some prior reports in nonhuman primates (Barrese et al. 2013; Sponheim et al. 2021). Even when array yields are low, decoding SNR can still be high due to the information contained in spike band power features (e.g., participant T6) (Nason et al. 2020), and incorporating spike band power features is now standard practice (Willett et al. 2023; Card et al. 2024; Gusman et al. 2025). All arrays except two yielded functional movement intention signals, and nearly all (11 of 14) did so throughout the duration of study participation. Array recordings generally showed higher decoding SNR over the most recent 10 years compared to previous arrays (but see T11 Lateral; Figure 3E), possibly reflecting process improvements over time. Neural tuning stability was also reasonably high in many arrays (correlation > 0.6 for day pairs within a 1-month time span). Good tuning stability allows decoders calibrated on a previous day to be used on a new day, reducing or eliminating the need for interrupting the user to recalibrate (Jarosiewicz et al. 2015; Sussillo et al. 2016; Wilson et al. 2023; Fan et al. 2023; Hosman et al. 2023; Card et al. 2024)

Despite favorable results overall, some arrays did fail to consistently produce useful decoding signals over time (3 of 14 arrays evaluated for decoding; T2, T3, T11 Lateral; Figure 3A), experienced large declines in yield within the first year (2 of 19; S2, S3; Figure 1A) or had consistently low yield (4 of 19; S1, T3, T6, T11 Lateral; Figure 1A). Additionally, there was substantial variability in decoding quality that was not fully accounted for by yield (Figure 3F). While some top performing arrays provided movement intention signals that enabled directional decoding comparable to able-bodied computer mouse movements, there was a wide range of decoding SNRs, and adding more electrodes to low performing arrays was not projected to enable top performance within the range of ~1000 electrodes. To ensure consistently high decoding SNR, it may be necessary to discover the underlying factors driving variability in signal quality, which could include disease state, underlying injury, implant location, implant technique, variability in device manufacture, and foreign body response.

It has been suggested that very late-stage ALS may lead to motor cortical signal loss (Lawyer and Netsky 1953; Eisen and Weber 2001; Vansteensel et al. 2024). This study included six participants living at the time with ALS (A1, T1, T3, T6, T7, T9), although none reached a completely locked-in state. For these participants, array yield was relatively high in some cases (A1, T1, T7, T9) and low in others (T3, T6), with no clear relationship to their remaining ability to move, speak, or use their eyes for communication. Decoding SNR could not be assessed in A1 and T1 (lack of stored high-bandwidth recordings); dSNR was poor in T3 (with low yield) but did not appear to decline over time in participants T6, T7 and T9. Although these dSNR and array yield measurements are somewhat variable, they appear to be consistent with the broader study group.

Implant location may also affect decoding SNR if the hand area of motor cortex contains tuning “hot spots” for directional cursor movement. If so, more precise presurgical neuroimaging or broader coverage of relevant areas of motor cortex could improve decoding SNR. Additionally, variable foreign body response could be a factor, since silicon microelectrode arrays are known to cause tissue disruption in animal models (Nolta et al. 2015) and neural connectivity can decline even while neurons are spared (Gregory et al. 2023; Chen et al. 2024). Surgical technique may also affect array performance, potentially via minimization of tissue disruption and foreign body response.

Finally, it is important to note that arrays included in this study spanned 15 years of varying manufacturing processes, which would have contributed to some of the variability we observed. Also, arrays implanted in early trial participants used arrays with electrodes 1.0 mm in length (S1, S2, A1, T1, T3, T6) whereas later electrodes were 1.5 mm in length (S3, T2, T5, T7 through T11); combined with individual differences in cortical thickness, this would be expected to introduce variability regarding which cortical layers were recorded.

Data collected over decades of human intracortical BCI research has demonstrated the growing potential of intracortical BCIs as a viable assistive technology, and many commercial efforts are currently underway to develop novel recording devices with improved durability and greater number of electrodes (Musk and Neuralink 2019; Sahasrabuddhe et al. 2021; Hettick et al. 2022). As these new recording technologies become available, progress toward more performant BCI systems will require standardized benchmarks with which to compare devices. For cursor control applications, a variety of performance metrics have been applied, including Fitts bit rate, achieved bit rate, typing rate, target acquisition time and path length (Hochberg et al. 2006; Simeral et al. 2011; Gilja et al. 2015; Jarosiewicz et al. 2015; Nuyujukian et al. 2015; Pandarinath et al. 2017; Brandman et al. 2018). However, these metrics are sensitive to task parameters (e.g., target hold time or click decoding latency) and decoding algorithms, which vary widely across studies and have rarely been tuned for optimal comparative performance. Here, we used a decoding SNR metric that quantifies the amount of linear movement direction information present in a neural population, independent of decoding algorithm and task parameters, enabling a meaningful comparison across 10 years of ever-evolving research session designs. This metric could be used to compare devices aiming to enable BCI cursor control, and variants of this metric may enable comparison across other domains (e.g., speech), serving as an informative complement to clinical outcomes assessments that measure the combined efficacy of a particular device and neural decoding approach in restoring useful function.

## Data Availability

All data required to reproduce these results will be publicly released upon acceptance in a peer-reviewed journal.

## Acknowledgements

We thank participants S1, S2, S3, A1,T1, T2, T3, T6, T7, T8, T9, T10, T5, T11 and their care partners for their generously volunteered time and effort as part of our BrainGate pilot clinical trials, and Beverly Davis, Kathy Yu, Sandrin Kosasih, Beth Travers, David Rosler, and Maryam Masood for administrative support. From June 2004 to May 2009, the BrainGate pilot clinical trials were sponsored and directed by Cyberkinetics Neurotechnology Systems. Since 2009, those original clinical trials and the subsequent trial (“BrainGate2”) have been conducted by academic institutions with grant support from the NIH, VA, and charitable foundations.

## Funding

This work was supported by Office of Research and Development, Department of Veterans Affairs (N2864C, N9228C, A4820R, A2295R, B6453R, B6459L, A6779L, P1155R, A2827R, A3803R), NIH NIDCD (R01DC009899), NIH NIDCD (U01DC017844), NIH NIDCD (R01DC014034), NIH NIDCD (U01DC019430), NIH NICHD-NCMRR (N01HD53403), NIH NICHD-NCMRR (N01HD10018), NIH NICHD-NCMRR (R01HD077220), NIH NINDS, (U01NS098968), NIH NINDS (U01NS062092), NIH NINDS (UH2NS095548), NIH NINDS (U01NS123101), NIH NICHD (RC1HD063931), Simons Foundation (543,045), Howard Hughes Medical Institute, Doris Duke Charitable Foundation, Conquer Paralysis Now (004698), AHA (19CSLOI34780000), ALS Association (20-MALS-553), Larry and Pamela Garlick, Samuel and Betsy Reeves, MGH-Deane Institute, The Executive Committee on Research (ECOR) of Massachusetts General Hospital, Wu Tsai Neurosciences Institute, Bio-X Institute at Stanford, Robert J. and Nancy D. Carney Institute for Brain Science, Brown University School of Engineering, Brown University Office of the Vice President for Research

## Author Contributions

FRW conceived the study design and approach. NVH and FRW wrote the manuscript and led the analysis, interpretation and visualization of the data. ES and JDS conducted preliminary data analyses which influenced the approach taken in this study. JPD led the initial academic-startup ‘BrainGate’ collaborative group and the clinical trial at Cyberkinetics, of which he was co-founder and CSO, which included the preclinical and early clinical studies. He provided review and guidance of the data acquisition, analyses and interpretation in subsequent studies. LRH is the sponsor-investigator of the multisite BrainGate2 pilot clinical trial and proposed the data collection approach that would enable cross-participant analyses. JDS was responsible for standardized data formatting and storage which enabled this work. The study was supervised and guided by FRW. All authors reviewed and edited the manuscript. The BrainGate Consortium constitutes all those appointed at academic or VA institutions (including, but not limited to Brown University, Case Western Reserve University, Massachusetts General Hospital, Stanford University, and VA Cleveland and VA Providence) who directly enabled or participated in data collection for the 14 participants included in this study (including planning and performing array placement surgeries, developing necessary software infrastructure, collecting neural recordings, designing research sessions, and supervising and guiding site related activities):

Abidemi Ajiboye, Abraham Caplan, Alex Acosta, Alisa Levin, Ammar Shaikhouni, Anastasia Kapitonava, Andrew Cornwell, Anish Sarma, Anisha Rastogi, Arto Nurmikko, Beata Jarosiewicz, Benjamin Shanahan, Benjamin Walter, Benyamin Meschede-Krasa, Beth Travers, Blaise Yvert, Bradley Buchbinder, Brandon King, Brian Franco, Brian Murphy, Brianna Karpowicz, Brittany Sorice, Carlos Vargas-Irwin, Carol Grant, Chaofei Fan, Chethan Pandarinath, Christine Blabe, Claire Nicolas, Damien Lesenfants, Daniel Bacher, Daniel Milstein, Daniel Rubin, Daniel Thengone, Daniel Young, Darrel Deo, David Brandman, David Chen, David Rosler, Dawn Taylor, Dimitra Blana, Donald Avansino, Douglas Crowder, Ed Chadwick, Emad Eskandar, Emery Brown, Erik Sudderth, Erin Kunz, Erin Gallivan Oakley, Etsub Berhanu, Ewina Pun, Foram Kamdar, Gerhard Friehs, Guy Wilson, Jacob Donoghue, Jacob Gusman, Jacqueline Hynes, Jad Saab, Jaimie Henderson, Jannis Brea, János Perge, Jason Pacheco, Jean-Baptiste Eichenlaub, Jennifer Sweet, Jeremy Shefner, Jessica Abreu, Jessica Feldman, Jessica Kelemen, Jie Liu, John Ciancibello, John Mislow, Jon Mukand, Jonathan Miller, Joris Lambrecht, Jose Albites-Sanabria, Justin Jude, Kaitlin Wilcoxen, Katherine Centrella Newell, Kathryn Tringale, Krishna Shenoy, Kristi Emerson, Kushaal Rao, Laurie Barefoot, Lucille Panagos, Marco Vilela, Marguerite Bowker, Mark Homer, Maryam Masood, Maryam Saleh, Matthew Harrison, Matthew Willsey, Michael Black, Michael Burkhart, Michael Keith, Mijail Serruya, Naryan Murthy, Natalie Herrick, Nicholas Schmansky, Nicolas Masse, Nir Even-Chen, Nishal Shah, Paul Nuyujukian, Paymon Rezaii, Rekha Crawford, Richard Penn, Robert Kirsch, Ronnie Gross, Rose Marujo, Sergey Stavisky, Shane Allcroft, Shaomin Zhang, Sharlene Flesher, Shaul Druckmann, Stefan Lutschg Espinosa, Stephen Mernoff, Stephen Ryu, Sung-Phil Kim, Susan Fasoli, Sydney Cash, Tanya Lewis, Thomas Hosman, Tomislav Milekovic, Tyler Singer-Clark, Vamsi Chavakula, Vikash Gilja, Wasim Malik, William Memberg, Wilson Truccolo, Ziv Williams

## Competing Interests

The MGH Translational Research Center has a clinical research support agreement (CRSA) with Ability Neurotech, Axoft, Neuralink, Neurobionics, Paradromics, Reach Neuro, Precision Neuro, and Synchron, for which LRH provides consultative input. LRH is a non-compensated member of the Board of Directors of a nonprofit assistive communication device technology foundation (Speak Your Mind Foundation). Mass General Brigham (MGB) is convening the Implantable Brain-Computer Interface Collaborative Community (iBCI-CC); charitable gift agreements to MGB, including those received to date from Paradromics, Synchron, Precision Neuro, Neuralink, and Blackrock Neurotech, support the iBCI-CC, for which LRH provides effort.

FRW is an inventor on intellectual property licensed by Stanford University to Blackrock Neurotech and Neuralink Corp.

JPD is a Director and shareholder in Pathmaker Neurosystems and a shareholder in Beacon Biosignals.

All other authors have no competing interests.

## Supplementary Figures

**Supplementary Figure 1.**
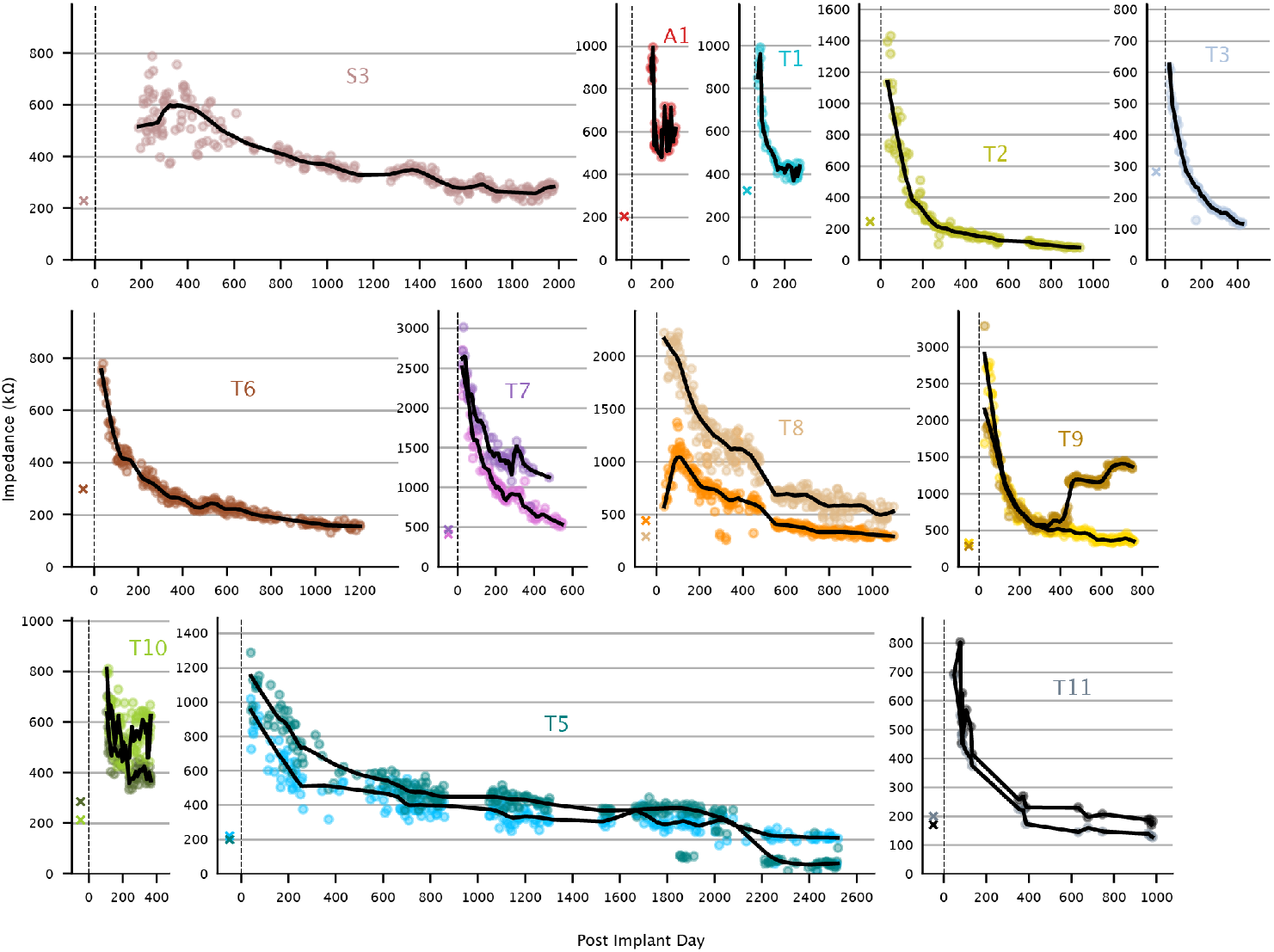
Median impedance. Median impedance measurements for 18 of 20 arrays. Points denoted with an X indicate the (manufacturer provided) pre-implant median impedance measurements. Impedance measurements were not recorded for Participant S1, and there were limited recordings for participant S2 so these arrays were excluded.

**Supplementary Figure 2.**
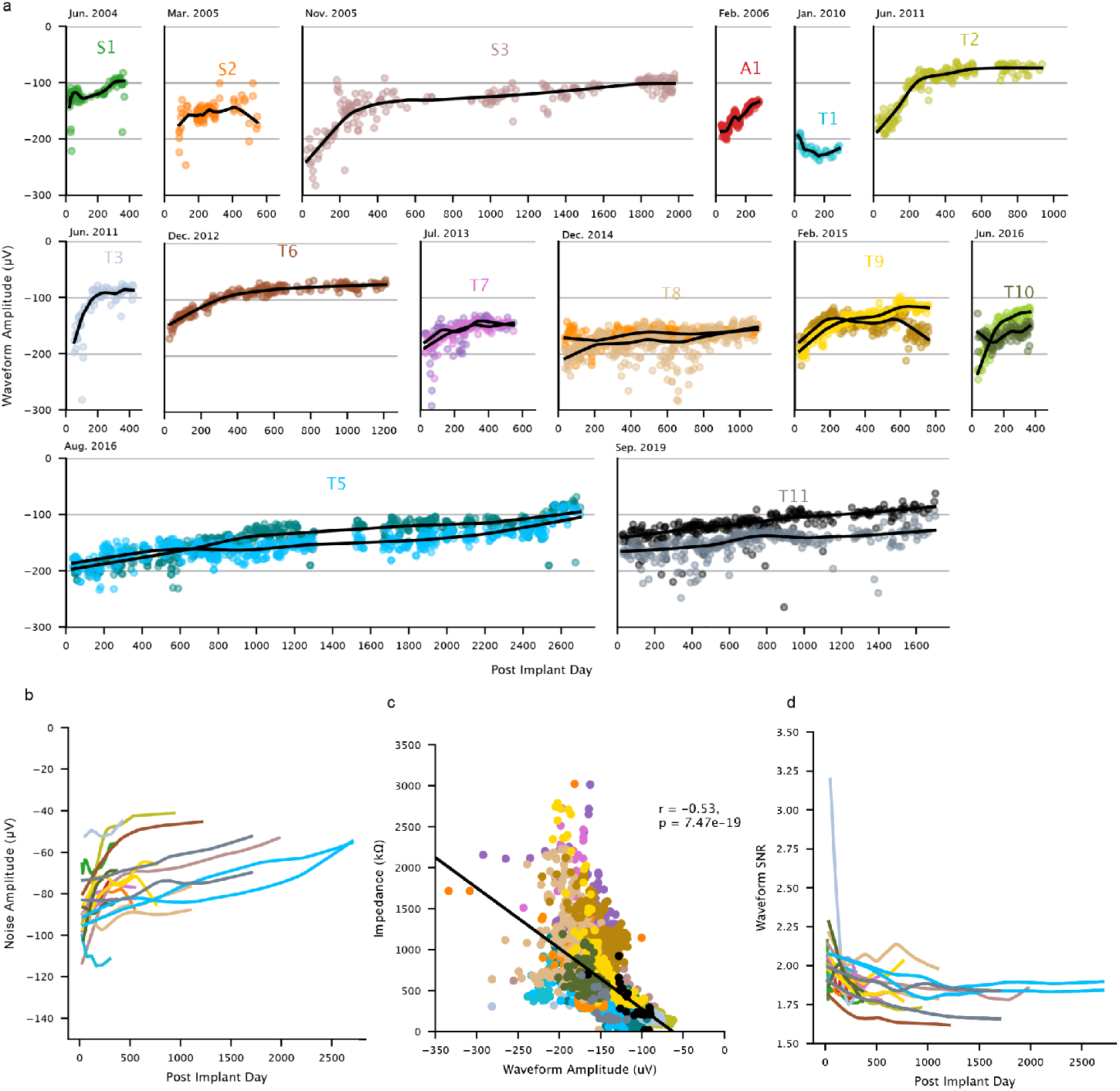
Spike waveform signal and noise. **(A)** Waveform amplitudes over time for each array. Using a −4.5 times robust standard deviation threshold, spike waveforms were extracted and averaged for each electrode. The minimum (negative) voltage was then taken as the waveform amplitude for each channel. Each point represents the median waveform amplitude for electrodes detecting a threshold crossing rate greater than 2 Hz in an array. A single neural recording from each session was used. LOWESS curves are shown in black. Waveform amplitudes generally decreased over time. **(B)** Noise amplitude LOWESS curves for each array. Noise amplitude was estimated as −3.0 x RMS of the voltage signal. Noise amplitudes generally decreased over time. **(C)** Waveform amplitude is inversely correlated with impedance (r: −.53, *p*<.001), consistent with insulation degradation. Each point represents a single session’s median impedance and waveform amplitude values. **(D)** Waveform SNR. Waveform SNR was calculated as the ratio of waveform amplitude and noise amplitude. SNR remained relatively constant over time after an initial decline, indicating that waveform and noise amplitudes decreased at similar rates and did not appear to affect the ability to identify spikes up to 7+ years post implant.

**Supplementary Figure 3.**
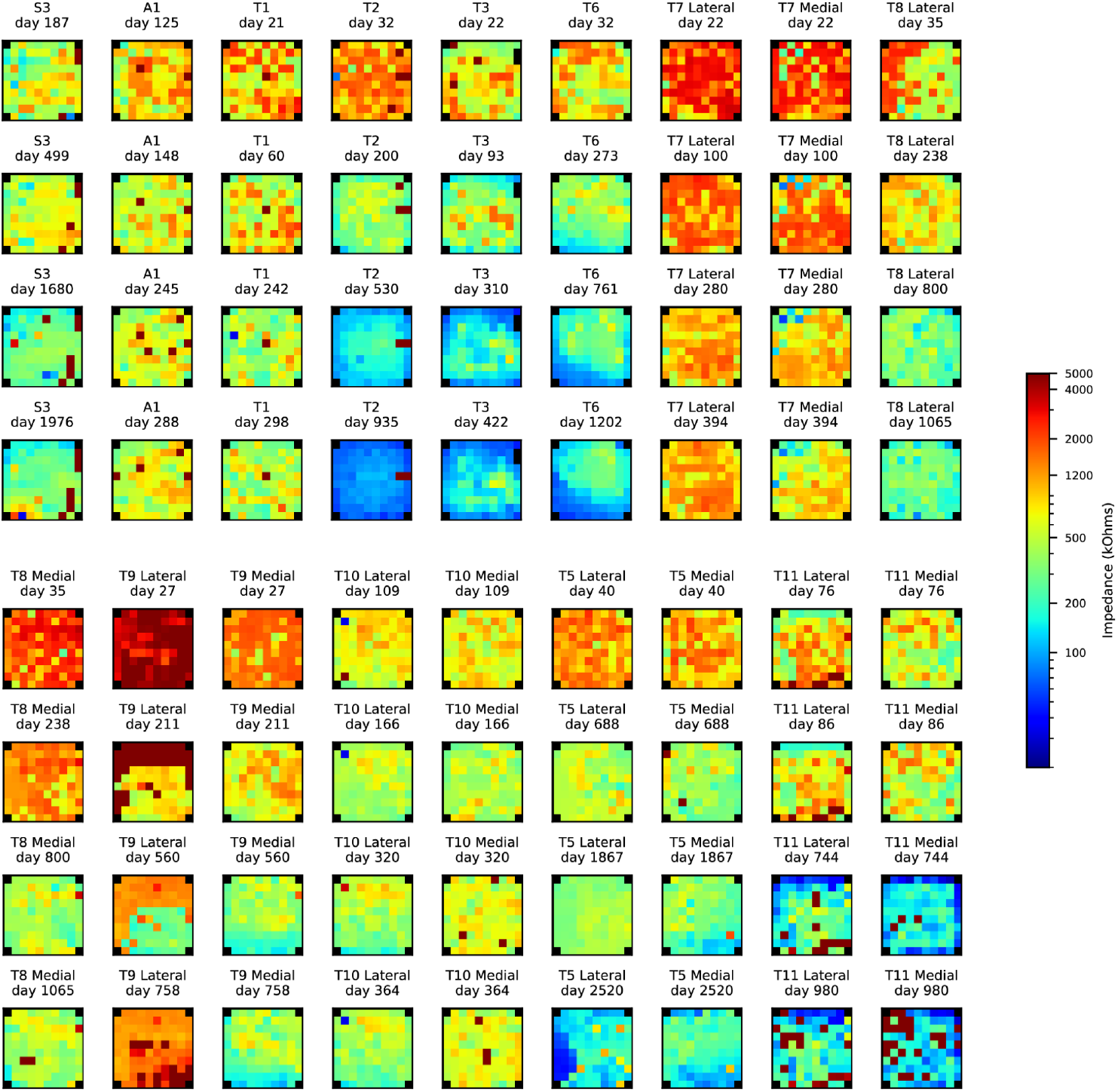
Impedance heatmaps. Impedance measurements for four sessions included in the impedance dataset for each array (see Supplementary Methods 2.3). Each individual square within an array denotes the impedance measured on that electrode; the four corner electrodes were not wired for recording. On some arrays, impedance appears to decline more sharply along the edges of the array.

**Supplementary Figure 4.**
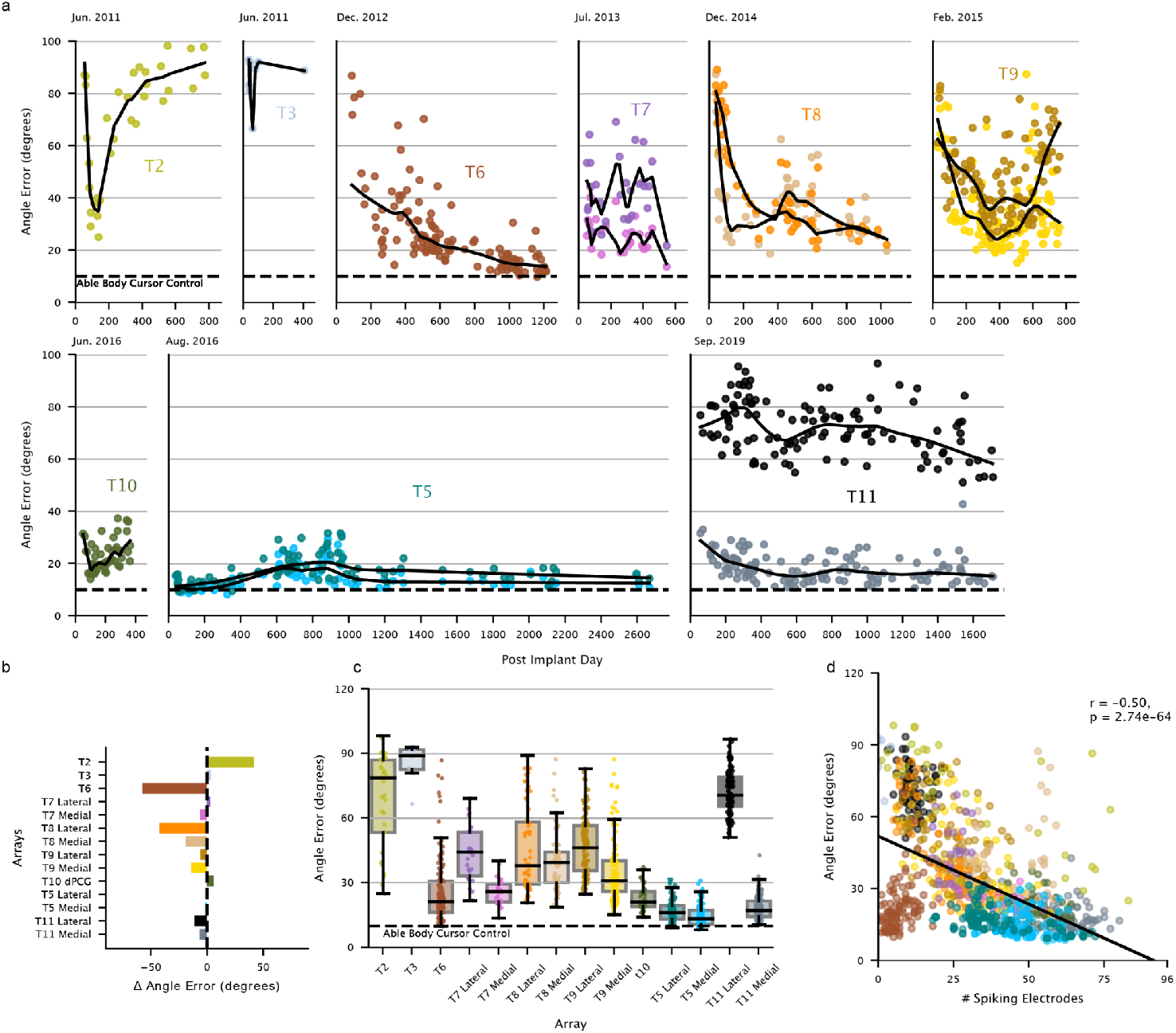
Angular error. Angular error is calculated as the absolute angle difference between the vector from cursor position to target position and the decoded vector. A 300 ms window of activity per trial was used to evaluate angular error, as opposed to an instantaneous angular error metric. Angular error ranges from 0 degrees (decoded vector is the same as inferred intended directional vector to 180 degrees (decoded vector is pointed directly opposite of the inferred intended directional vector. **(A)** Angular error over time. Each point represents an angular error value for one session. LOWESS curves are shown in black for each array. **(B)** Overall deltas in angular error. Deltas were calculated by taking the mean angular error in sessions recorded during the first 3 months post implant and subtracting the mean of angular error recorded during the last 3 months of recording. **(C)** Angular error distributions. Arrays are displayed in implant order. The dotted line indicates mean angular error across 9 able body participants performing the cursor task **(D)** Angular error and array yield are inversely correlated overall (r: −0.50, *p* < .001), although yield does not fully explain differences in decoding performance across arrays (same as in Figure 3f).

**Supplementary Figure 5.**
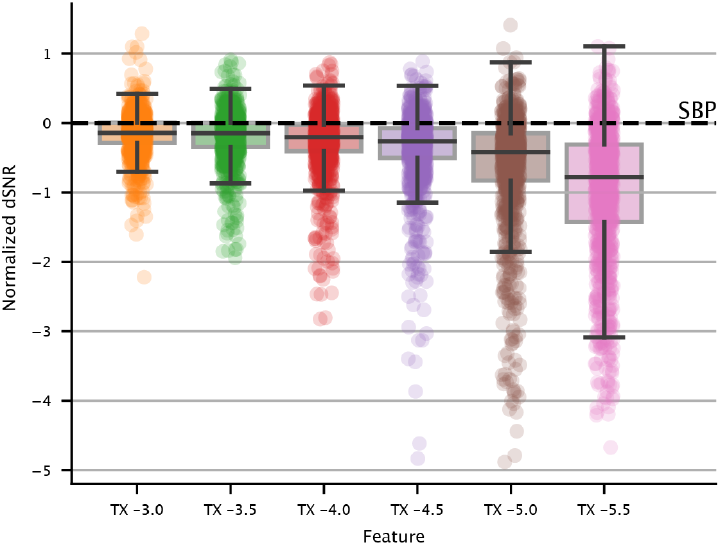
Neural features. Distribution of normalized dSNR values. dSNR values for threshold crossings extracted with thresholds from a range of threshold multipliers are shown relative to dSNR values for spike band power features. As the threshold multiplier approached zero, decoding performance increased.

## Supplementary Methods

### 1 Research procedures

### 1 Experimental procedures

#### 1.1 BrainGate and BrainGate2 Clinical Trials

The BrainGate feasibility study (ClinicalTrials.gov Identifier: NCT00912041) is the largest and longest-running clinical trial of an implanted BCI. From June 2004 to May 2009, there were two IDEs: one enrolling individuals with spinal cord injury or brainstem stroke, and another that enrolled individuals with motor neuron disease. In 2009, clinical trial sponsorship transitioned from Cyberkinetics, Inc to Massachusetts General Hospital and a second generation trial “BrainGate2” was initiated, inclusive of people with diagnoses of cervical spinal cord injury, brainstem stroke, muscular dystrophy, or motor neuron disease.

#### 1.2 Study participants

This study includes data from the first 14 participants in the BrainGate and BrainGate2 pilot clinical trials (see Supplementary Table 1, modified from Rubin et al. 2023), who each gave informed consent prior to any research procedures. Participants were enrolled in the BrainGate feasibility studies (BrainGate2 ClincialTrials.gov Identifier: NCT00912041) which is performed under an Investigational Device Exemption (IDE) from the US Food and Drug Administration and approved by the Mass General Brigham Institutional Review Board (Protocol #: 2009P000505; CAUTION: Investigational Device. Limited by Federal Law to Investigational Use). Permission was also granted by the Stanford University Institutional Review Board (IRB; protocol #20804), the Mass General Brigham IRB (protocol #2009P000505), Case Western Reserve University IRB, and the Providence VA Healthcare IRB. All research was performed in accordance with relevant guidelines and regulations.

**Supplementary Table 1.** Arrays and sessions analyzed.

| Participant | Gender | Status | Time from injury/<br>diagnosis to<br>implant (y) | Implant Date | No. of arrays | Electrode Length (mm) | Total Implant Days | Etiology |
| --- | --- | --- | --- | --- | --- | --- | --- | --- |
| S1 | M | Explanted | 3 | 6/2004 | 1 | 1.0 | 484 | C4 SCI, AIS-A |
| S2 | M | Explanted | 6 | 3/2005 | 1 | 1.0 | 790 | C4 SCI, AIS-A |
| S3 | F | Explanted | 10 | 11/2005 | 1 | 1.5 | 1994 | Pontine Stroke |
| A1 | M | Deceased | 7 | 2/2006 | 1 | 1.0 | 296 | ALS |
| T1 | F | Deceased | 7 | 1/2010 | 1 | 1.0 | 307 | ALS |
| T2 | M | Explanted | 5 | 6/2011 | 1 | 1.5 | 950 | Pontine Stroke |
| T3 | M | Deceased | 2 | 6/2011 | 1 | 1.0 | 440 | ALS |
| T6 | F | Explanted | 7 | 12/2012 | 1 | 1.0 | 1216 | ALS, ALSFRS -R 16 |
| T7 | M | Deceased | 3 | 7/2013 | 2 | 1.5 | 552 | ALS, ALSFRS -R 17 |
| T8 | M | Deceased | 8 | 12/2014 | 2 | 1.5 | 1104 | C4 SCI, AIS-A |
| T9 | M | Deceased | 4 | 2/2015 | 2 | 1.5 | 759 | ALS |
| T10 | M | Explanted | 9 | 6/2016 | 2* | 1.5 | 518 | C4 SCI, AIS-A |
| T5 | M | Deceased | 9 | 8/2016 | 2 | 1.5 | 2780 | C4 SCI, AIS-C |
| T11 | M | Enrolled | 11 | 9/2019 | 2 | 1.5 | ** | C4 SCI, AIS-B |

One or two 96 channel intracortical microelectrode arrays (Cyberkinetics Neurotechnology Systems (S1-S3, A1); Blackrock Neurotech (T1-T11)) were placed in the cortex of 14 participants as part of the BrainGate and BrainGate2 clinical trials. *In T10, one array was placed in middle frontal gyrus. This array was included in yield analyses but not decoding analyses (since it was placed in a different brain area that is not comparable to hand knob). **Participant T11 is currently still enrolled and participating in research sessions. These analyses included T11 implant days 0-1710.

#### 1.3 Electrode properties

All arrays included in this study had platinum metallization of the electrode tip. BrainGate participants after those reported here have instead used iridium oxide arrays which have shown superior yield in non-human primates (Sponheim et al. 2021).

Electrode length was either 1.0 mm or 1.5 mm depending on the participant. At the beginning of the trial, participants with ALS were provided with 1.0 mm arrays even when the transition to 1.5 mm arrays had been made for participants with SCI or brainstem stroke. This was informed by the concern that purposefully using 1.5 mm electrodes whose tips might rest in cortical layer V, where the Betz corticomotoneuronal cells are located, might be less informative (due to Betz cell loss in ALS) than aiming for more superficial layers. With experience, we recognized that variations in the thickness of the arachnoid and subarachnoid space, as well as imperfect ability to sit the array perfectly tangential to the (curved) cortical surface made it likely that array tips were often sitting in either superficial layer V or above even with 1.5mm electrodes.

#### 1.4 Neural signal processing

High-bandwidth neural signals were recorded from silicon microelectrode arrays using Blackrock Neurotech’s NeuroPort system. Signals were analog filtered (4th order Butterworth with corners at 0.3 Hz to 7.5 kHz) and digitized at 30 kHz. For this study, all additional signal processing was performed offline in order to standardize the signal processing pipeline across participants, sessions, and experimental designs.

First, 30 kHz neural recordings (Blackrock Neurotech .ns5 files) were decimated to 15 kHz and bandpass filtered between 250 Hz to 5000 Hz using a 4th order Butterworth filter. Next, denoising was applied using LRR (Young et al. 2018; Musial et al. 2002) LRR computes a unique reference signal for each channel. The LRR coefficients are calculated using linear regression to predict a channel’s activity based on the activity of all other channels in the array. These coefficients are then applied to the recording, generating an LRR signal which is then subtracted from the channel’s recorded voltage.

Two classes of features were then computed: binned spike band power and binned threshold crossings. Unsorted threshold crossings and spike band power are commonly used measurements of local spiking activity that enable comparable (or better) decoding performance as compared to sorted spikes (Chestek et al. 2011; Christie et al. 2015; Stark and Abeles 2007; Nason et al. 2020). Binned spike band power was computed by taking the sum of squared voltages observed in 10 ms bins. Binned threshold counts were computed by counting the number of times the filtered voltage time series crossed the amplitude threshold in a 10 ms period. Electrode specific thresholds were calculated using a multiplier (−3.0, −3.5, −4.0, −4.5, −5.0, and −5.5) times the standard deviation of the voltage signal for each electrode. “Robust” thresholds were calculated using the same multipliers but applied to the scaled median absolute deviation (MAD × 1.4826) of the voltage signal for each electrode. The scale factor of 1.4826 was used to make the MAD equivalent to the standard deviation for normally distributed data. In addition to binned threshold counts, spike “snippets” were saved for all threshold crossing events. A snippet consisted of a 15 kHz filtered voltage trace with 10 samples before the detected crossing, and 22 samples including and after the detected crossing. Snippet data captured ~2.1 ms waveforms of neural spikes. Electrode specific thresholds and LRR filter coefficients were computed from and applied to each .ns5 recording independently.

**Supplementary Methods Figure 2.**
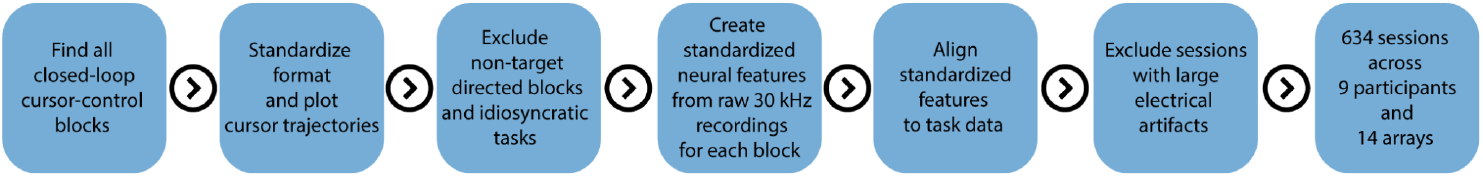
Analysis pipeline for the decoding dataset.

#### 1.5 Overview of data collection sessions

Neural data were recorded from each participant in sessions lasting approximately 2-5 hours, typically occurring twice or more per week. Sessions could be cancelled or ended early at the participant’s request. Participants were seated in a wheelchair or bed facing a computer monitor. Each session was divided into blocks of uninterrupted trials for a specific task. Participants were encouraged to rest between blocks, with block durations typically ranging from 1 to 20 minutes based on their preference. The goal of developing and testing new neural decoding architectures resulted in a wide variety of tasks and daily research block designs across sessions and participants.

##### 1.5.1 Recording sessions used to estimate array yield

Neural data from 2,319 sessions across 14 participants were evaluated in Figure 2, Supplementary Figure 1, and Supplementary Figure 2 to estimate array yield (“yield dataset”). The yield dataset consists of a single high-bandwidth recording from each session (.ns5 file), used to derive threshold crossing rates and spike waveform snippets. A single recording from each session was programmatically selected based on its duration (the shortest recording greater than 5 minutes was chosen; if no recordings were longer than 5 minutes, the longest recording was chosen). Research task or device settings (e.g., headstage used, reference wire used) were not considered. Raster plots and spike panels were generated for all candidate sessions using a robust threshold with a −4.5 multiplier (see Supplementary Methods 1.3) and manually inspected. Sessions with evident electrical artifacts were discarded. A total of 2,486 candidate sessions were evaluated and 2,319 were included in the study (see Supplementary Methods Figure 1 Below).

**Supplementary Methods Figure 1.**
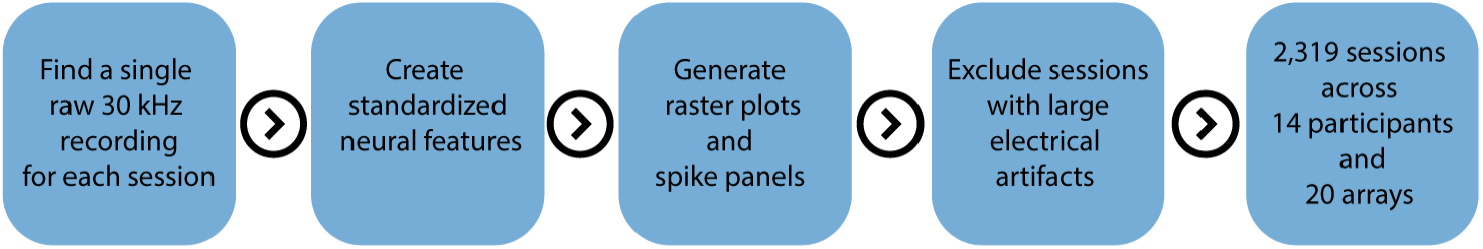
Analysis pipeline for the yield dataset.

##### 1.5.2 Recording sessions used to estimate decoding SNR

Neural and BCI cursor control task data from 634 sessions across 14 arrays and 9 participants were evaluated in Figure 3 to estimate decoding SNR (“decoding dataset”). Relative to the “yield dataset” above the decoding dataset contains a more limited number of sessions and participants, as not all sessions included closed-loop BCI cursor control, and for the first 5 participants high-bandwidth neural data (.ns5 files) were either not recorded during cursor control (S1, S2, A1), not recorded for the first year post-implant (S3), or could not be consistently aligned to cursor task data (T1). For each session, binned threshold crossings, binned spike band power, target positions, and cursor positions were extracted from all candidate closed-loop cursor control blocks collected within that session. Candidate closed-loop cursor control blocks were identified as blocks involving goal-directed movement towards explicit targets (eg. tasks such as radial-8, fitts, grid task) based on task metadata and documented session notes. Cursor trajectories were plotted and non-target directed blocks were excluded, as well as idiosyncratic tasks, and blocks where poor closed-loop decoder performance was likely to prevent consistent movement intention behavior from participants (e.g., cursor stuck in a corner of the screen for long durations due to noise or errors in novel/experimental decoding pipelines).

Binned threshold crossings and spike band power were calculated using 10, 20, 50 or 100 ms bins from high bandwidth recordings (see Supplementary methods 1.4) and aligned to target positions and cursor positions measured at the same resolution (bin width varied depending on the participant and session, and were the same bin widths used at the time of data collection). Bin widths greater than 20 were only used for T2 and T3. Raster plots were generated from ns5 derived features and blocks with excessive electrical noise were discarded. Data from individual blocks within a session were then concatenated into a single dataset for each research session. A total of 712 candidate cursor control sessions were evaluated, and 634 were included in this study (see Supplementary Methods Figure 2).

### 1.5 Able-bodied mouse movements

Able-bodied mouse movements were recorded from 9 volunteers who engaged in a task designed to assess their cursor control accuracy when using an optical computer mouse (Dell MS111) and linux computer (Ubuntu 22.04). This study was reviewed and approved by Stanford University Institutional Review Board under protocol number 68028. Each participant was seated in an office chair facing a computer monitor that displayed a ring of targets surrounding a central target. Data collection occurred during a single session, which comprised three blocks of 100 trials each. Participants were instructed to move the cursor from the center of the ring to a designated target as quickly, accurately, and consistently as possible. During each trial, a single target in the ring would illuminate in green, prompting participants to initiate their movement. This typically resulted in an initial “push” towards the target, followed by smaller corrective movements towards the target. After hovering the cursor above the target for 500 ms, the central target would turn green, signaling participants to return the cursor to the center in preparation for the subsequent trial.

## 2 Longevity metrics

### 2.1 Spiking electrodes and array yield

The number of spiking electrodes was calculated for each session included in the yield dataset (defined in 1.5.1). Linear regression referencing and a robust −4.5 threshold was used to extract threshold crossings (see 1.4). To obtain a robust measure of threshold crossing rates, we segmented the data into 10 second intervals and computed the threshold crossing rate for each segment. The median threshold crossing rate was then determined to mitigate the influence of outliers. Electrodes that detected a median threshold crossing rate greater than 2 Hz were considered to be “spiking” and contributed to the overall spiking electrode count of the array. A 2 Hz threshold was based on manual inspection of spike waveforms and is a conservative measure that aligned with our perception of which electrodes were active on a spike panel visualization. Array yield was then calculated as the percentage of spiking electrodes in the array.

### 2.2 Spike waveform signal and noise

Neural spike waveform amplitude and noise amplitude were calculated for each session included in the yield dataset (defined in 1.5.1). Spike waveform snippets were extracted (as described in 1.3, with linear regression referencing and a −4.5 robust threshold) and for each spiking electrode, a mean spike waveform snippet was calculated by averaging all waveform snippets over time. The minimum voltage value for the mean snippet was then denoted as the spike waveform amplitude for that electrode. The median spike waveform amplitude was then used to compare waveform amplitudes over the recording duration of the array (Supplementary Figure 2). To identify noise amplitudes over time, we took the mean robust threshold value across spiking electrodes when using a −3.0 multiplier (see 1.3 for thresholding details).

### 2.3 Impedance

Array impedance values were calculated for all sessions in the yield dataset (defined in 1.5.1) that had acompanying recorded impedance values. As not all sessions included impedance data collection, this dataset consisted of a more limited number of sessions and participants (1,646 of 2,319 total sessions). Impedance measurements were not recorded for participant S1, and a limited number of sessions had impedance measurements for participant S2 (not allowing for a meaningful trend to be derived), so these arrays were excluded. Automated impedance measurements became available on implant day 125 for participant A1. For sessions with multiple impedance recordings, a single instance was used. Electrode impedance measurements were taken through the percutaneous pedestals using Blackrock pogo pin or thin film patient cables and Blackrock Neurotech’s Central software and hardware (1 kHz test frequency). The median impedance value across channels was taken for comparing impedance values over recording duration (Supplementary Figure 1). Electrode impedance measurements over 4000 kOhms were outliers indicating non-functioning electrode connections and were discarded in Supplementary Figure 1. To visualize impedance measurements relative to the array geometry in Supplementary Figure 3, impedances are shown for four days spanning the implant period: the first post-implant measurement, the last available impedance measurement (which was prior to the last trial day for some participants), and 2 measurements in between.

### 2.4 Decoding Signal-to-Noise Ratio (dSNR)

The decoding signal-to-noise ratio (dSNR) is a vector-based movement intention decoding metric decomposing offline linear decoder output into a signal component (pointing toward the target) and a noise component to calculate a decoding SNR. Variations of this metric have been applied to iBCI data for cursor control and finger movement decoding (Shah et al. 2024; Willett, Murphy, et al. 2017; Willsey et al. 2025)

#### 2.4.1 BCI dSNR

Decoding signal-to-noise ratio was evaluated for all closed loop cursor sessions included in the decoding dataset (defined in 1.5.2; this dataset includes all target-directed cursor control tasks such as radial 8, grid, Fitts, etc.). For each session, neural activity, cursor positions, and target positions were concatenated across multiple experimental blocks. Neural features were block-wise normalized by subtracting the median (for spike band power) or mean (for threshold crossing rates) to account for nonstationarities in neural activity. Additional normalization was performed on spike band power features by dividing by the robust standard deviation (1.4826 times the median absolute deviation) and capping values at 100.

To calculate a dSNR value for each session, we first calculated a 2 × 1 position error unit vector, 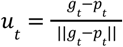, for each time bin *t*, where *g*_*t*_ is the target position and *p*_*t*_ is the cursor position. Bins where the distance to target ‖*g*_*t*_ − *p*_*t*_ ‖ was less than a participant specific distance threshold were excluded, as movement intention becomes ambiguous when the cursor is already close to the target. The remaining bins were divided into 5 cross-validation folds. For each fold, we trained a linear decoder to predict *u*_*t*_ as a linear function of the corresponding neural feature vector *f*_*t*_ :

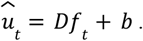

Here,

*D* is an 2 x N decoder matrix, *f*_*t*_ is an N x 1 feature vector, and *b* is a 2 × 1 bias term. We estimated *D* and *b* using ordinary least squares.

Because of regression dilution, noise in *f*_*t*_ causes *D* to shrink, and the linear decoder outputs are biased to be smaller than the unit vector targets *u*_*t*_. To correct for this effect and force the decoder’s output to lie around the unit circle (which is helpful for visualization and estimation of SNR), we re-normalized *D* and *b* after training:

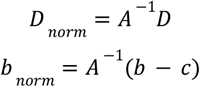

We estimated the 2 × 2 matrix *A* and the 2 × 1 column vector *c* by modeling how window-averaged decoder outputs from the training set related to the ground truth. Let *y*_*i*_ be the 2 × 1 average decoder output from a 300 ms window at the beginning of a trial *i* (beginning after a participant-specific reaction time). We then estimated *A* and *c* using ordinary least squares applied to the model:

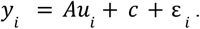

It then follows that if we would like *y*_*I*_ to match the scale of the ground truth vectors *u* _*i*_, we can subtract *c* and multiply by *A* ^−1^, which results in outputs that match *u* on average:

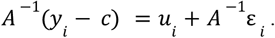

Finally, we evaluated the SNR of the held-out decoder predictions across all folds. Let *y*_*norm,i*_ be the output of the normalized decoder for trial *i*, averaged across a 300 ms window at the beginning of the trial. We fit the following linear model to *y*_*norm,i*_ :

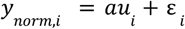

Here, *a* is a scalar parameter that captures the scale of the decoder outputs (if normalization was successful, *a* will be near 1) and *ε*_*i*_ is a 2 × 1 gaussian noise vector. The *a* parameter was fit using ordinary least squares. To estimate the magnitude of decoding error, observed model errors *ϵ*_*i*_ = *ay*_norm,i_ − *u*_*I*_ were computed and combined over the X and Y dimensions into a single vector. Finally, the standard deviation of *ϵ*_*i*_ was estimated in a robust way by multiplying the median absolute deviation by 1. 4826 (*σ* = 1. 4826 × median(|*ϵ*|)). dSNR was then calculated as 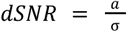.

To estimate session-specific reaction times without overfitting the reaction time, we employed an outer 10-fold cross-validation procedure. For each outer fold, reaction time was swept in 100 ms increments from 0 to 1000 and dSNR was estimated. The reaction time yielding the highest dSNR was then chosen and applied to generate window-averaged decoder outputs for all trials in the held-out test set. The window-averaged outputs across all test sets were then combined to estimate a final dSNR value per session (note that reaction times may have varied across folds and across sessions).

#### 2.4.2 Able-bodied mouse movements dSNR

To assess the decoding signal-to-noise ratio in able-bodied participants performing cursor movements (see 1.6) we used a modified dSNR metric. Cursor trajectories were binned by taking the median cursor position in 10 ms windows. Movement onset was defined as 30 ms prior to the cursor speed exceeding a threshold of 1 (arbitrary units). Evaluation windows captured the initial movement while excluding corrective movements after the initial “push” (see Supplementary Methods Figure 3). As speeds differed between participants, the evaluation window length used for the analysis period was participant specific and determined by sweeping window lengths from 160 ms to 260 ms to maximize dSNR. A movement vector was calculated between cursor positions at the end of the evaluation window and the start of the evaluation window. This vector was used in place of the decoder predicted point at target used for iBCI dSNR.

**Supplementary Methods Figure 3.**
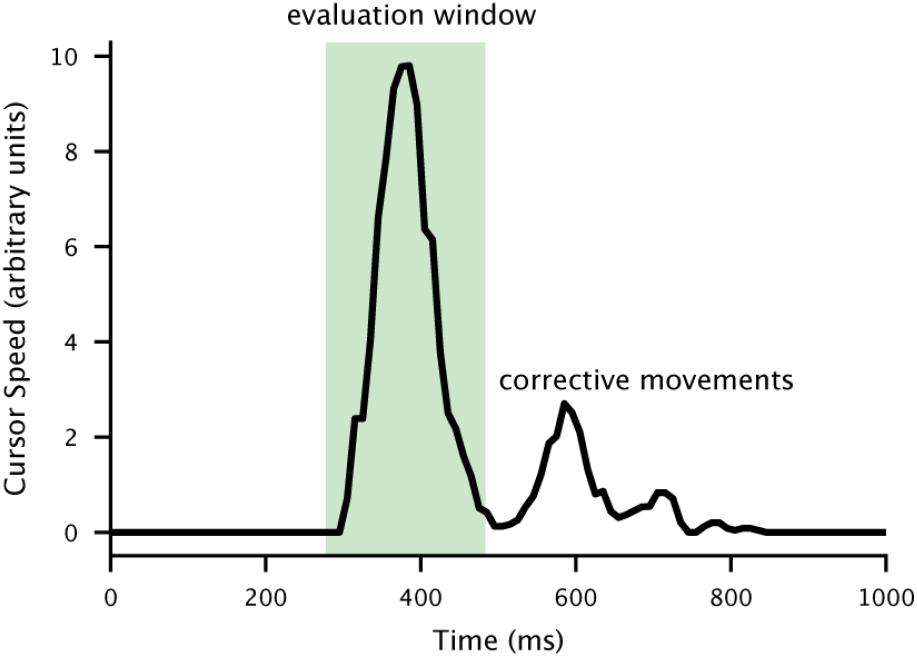
Mouse movement evaluation window.

### 2.5 Angular Error

Angular error is defined as the absolute angle difference between the inferred intended movement vector from cursor position to target position and the decoded vector. An angular error of 0° indicates perfect alignment between the intended and decoded movement directions, while an error of 180° indicates the decoded direction is opposite to the intended direction. Angular error complements dSNR by providing a more intuitive measure of directional accuracy that is directly interpretable in the context of cursor control tasks. While SNR captures both magnitude and direction, angular error specifically quantifies directional accuracy.

#### 2.5.1 iBCI angular error

Angular error was evaluated for all closed loop cursor sessions described in 1.5.2 using the same cross-validated decoding framework used for dSNR calculations. For each trial, the intended movement direction was defined as the unit vector from cursor position to target position during the 300ms evaluation window. The decoded direction was obtained from the linear decoder’s output on held-out test data. The mean angular error across trials was taken to be the angular error for that session.

#### 2.5.2 Able-bodied mouse movements angular error

For able-bodied mouse movement data, angular error was computed between the actual movement vector and the intended target direction for each trial. Using the same movement epochs identified for dSNR analysis (see 2.4.2), the angular error was calculated as the absolute angle between the movement vector and the unit vector pointing from initial cursor position to target.

### 2.6 Tuning Stability Metric

Neural tuning stability was evaluated for 11 of 14 arrays included in the decoding dataset (defined in 1.5.2). Three arrays (T2, T3, T11 Lateral) were discarded from this analysis, as they did not consistently provide meaningful movement intention decoding (dSNR <1) for the cursor tasks evaluated. Spike band power features were block-wise normalized by subtracting the median. Additional normalization was performed by dividing by the robust standard deviation (1.4826 times the median absolute deviation) and capping spike band power values at 100.

For each session, we estimated a linear model of neural tuning to intended movement direction:

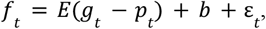

where *f*_t_ is an N x 1 neural feature vector at time step *t, E* is an N x 2 matrix of neural tuning coefficients, *g*_t_ is a 2 × 1 target position vector, *p*_t_ is a 2 × 1 cursor position vector, *b* is an N x 1 bias term, and *ε*_t_ is an N x 1 vector of Gaussian noise. *E* and *b* were estimated using robust regression to minimize the effects of any time steps containing electrical artifacts. We used the “robustfit” function in MATLAB (MATLAB R2024b, The MathWorks Inc) with bisquare weighting and a 4.685 tuning coefficient (Holland and Welsch 1977).

To obtain estimates of ‖*E*_x_ ‖ and ‖*E*_y_ ‖ (the columns of *E* corresponding to the *x* and *y* directions), which are used to compute across-day correlations, we used two-fold cross-validation to reduce bias. Otherwise, simply computing ‖*E*_x_ ‖ and ‖*E* _y_‖ from a linear model estimate of *E* results in estimates that are biased upwards (noise in the estimates inflates the magnitude). We fit separate linear encoding models *E*_1_ and *E*_2_ on two independent folds following the methods above, centered the columns of *E*_1_ and *E*_2_ by mean-subtracting each column, and then estimated ‖*E*_*x*_ ‖ as follows (and proceeding similarly for ‖*E* _*y*_‖):

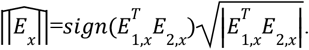

This estimate is implemented as cvOLS in the cvVectorStats library (https://github.com/fwillett/cvVectorStats), along with a demonstration of efficacy (testOLS).

The correlation across days in the x-direction (and identically for y) can then be computed as:

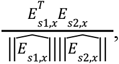

where *E*_*s*1,*x*_ is the column of x-direction coefficients for session 1, and *E*_*s*2,*x*_ is the column of x-direction coefficients for session 2. We averaged the x and y correlations to yield a single correlation value for each pair of sessions.

### 2.7 Electrode Scaling

Electrode scaling properties were assessed for 11 arrays providing meaningful movement intention decoding (dSNR > 1). To evaluate how decoding performance scales with electrode count, we randomly subsampled electrodes from 6 up to 96 and for each electrode count 500 random permutations were generated and 5 fold cross-validated dSNR was calculated using spike band power neural features (as described in 2.4.1). The mean dSNR across permutations was calculated for each electrode count, providing a scaling curve for each array. For participants with dual arrays, electrodes were pooled across both arrays and sample counts were extended to a combined 192 electrodes. dSNR scaled as an approximate logarithmic function of the number of electrodes included for decoding and linear extrapolation was used to estimate yields for electrode counts up to 1024.

## Notes

### Clinical Trial

NCT00912041

### Author Declarations

The IRB of Stanford University, IRB of Mass General Brigham, IRB of Case Western Reserve University and the IRB of Providence VA Healthcare gave ethical approval for this work

